# Risk stratification for the rapid pain progression phenotype in knee osteoarthritis using interpretable multimodal machine learning: Development in the Osteoarthritis Initiative and external evaluation in the Prospective Cohort of Osteoarthritis from A Coruña

**DOI:** 10.64898/2026.07.21.26357764

**Authors:** Francisco J. Blanco, Laura Martínez-Sotodosos, Natividad Oreiro, Laura Galindo, Jorge Vázquez-García, Diana M. Noriega-Cobo, Lucía Lourido, Rocío Paz-González, Patricia Quaranta, Valentina Calamia, Cristina Ruiz- Romero, Ignacio Rego-Pérez

**Author notes:** Both contributed equally to this paper (Francisco J. Blanco, Laura Martínez-Sotodosos). **Corresponding Address:** Instituto de Investigación Biomédica de A Coruña (INIBIC), Complexo Hospitalario Universitario de A Coruña (CHUAC), Sergas., As Xubias, 15006, A Coruña, Spain, and.

## Abstract

**Objective:** To develop an interpretable multimodal machine-learning model for risk stratification of the rapid pain progression phenotype in knee osteoarthritis and to evaluate its performance in the independent PROCOAC cohort.

**Methods:** An elastic-net logistic regression model was trained using Osteoarthritis Initiative (OAI) data. Rapid pain progression was defined over overlapping 24-month windows using normalized WOMAC pain. Harmonized clinical, genetic and proteomic candidates were evaluated, with feature selection by permutation importance. The frozen algorithm was tested in an OAI hold-out set and externally evaluated in PROCOAC. Logistic recalibration corrected prevalence shifts. Clinical utility was assessed by decision curve analysis.

**Results:** OAI comprised 2,934 individuals and 14,488 instances. Feature pruning reduced 159 candidates to a 19-variable clinical-genetic signature driven by Kellgren-Lawrence grade, localized knee pain, BMI and two genetic variants (rs73631790, rs9912678); no proteomic variable was retained. External testing in PROCOAC (582 individuals, 1609 instances) showed ROC-AUC 0.744 (95% CI 0.714 to 0.772) and PR-AUC 0.519. Following recalibration, the sensitive screening threshold yielded NPV 0.875 (95% CI 0.849 to 0.898) and sensitivity 0.804 (95% CI 0.760 to 0.844), whereas the high-specificity threshold achieved PPV 0.610 (95% CI 0.523 to 0.692) and specificity 0.941 (95% CI 0.924 to 0.954). Decision curve analysis showed positive net benefit at both thresholds, supporting a three-tier risk stratification framework.

**Conclusions:** This externally evaluated model identified patients at risk of rapid pain progression using an MRI-free clinical-genetic signature. Recalibrated thresholds may support risk-adapted monitoring, advanced imaging prioritization and trial enrichment.

## INTRODUCTION

Osteoarthritis (OA) is a complex, heterogeneous, whole-joint disease driven by the interplay of biomechanical stress, aging, metabolic dysfunction, low-grade inflammation, genetic susceptibility, and maladaptive tissue repair ^1–3^. Currently affecting over 595 million people worldwide, knee OA is one of the leading causes of global disability ^4^. Knee OA is particularly relevant as one of the most disabling phenotypes, frequently affecting mobility, independence, work participation and quality of life ^2–5^. Among all clinical manifestations, pain is the dominant symptom, and the main reason patients seek medical care ^3^.

Especially, pain severity in OA does not reliably correlate with radiographic joint space narrowing. This discordance highlights that symptoms are mediated by complex neurosensory and structural changes, such as bone marrow lesions and synovitis, often invisible on plain radiographs but detectable by magnetic resonance imaging (MRI) ^6–9^.

Longitudinal studies have shown heterogeneous pain trajectories in knee OA, with a subgroup of individuals experiencing clinically meaningful worsening associated with poorer outcomes, including an increased risk of total knee replacement ^10,11^. The rapid pain progression phenotype in OA (RPPOA) represents a phenotype of particular interest because it captures rapid symptom deterioration, potentially enabling earlier identification of high-risk individuals and improving the study of symptomatic progression in knee OA ^3,12^.

Despite the burden of OA pain, current therapeutic options remain limited. Standard management relies heavily on symptom reduction and late-stage joint replacement, as no disease-modifying OA drug (DMOAD) has yet achieved routine clinical approval ^13–15^. Identifying individuals at risk for RPPOA is therefore a clinical priority, as it would enable proactive monitoring, optimize the deployment of advanced imaging resources like MRI, and facilitate the enrichment of DMOAD clinical trials by safely isolating patients who are most likely to experience measurable progression ^3,12,16^.

Machine Learning (ML) provides a powerful framework for this type of stratified approach, capable of integrating multimodal clinical, demographic, and molecular variables to capture complex, non-linear prognostic patterns of knee OA ^17^. Recent studies have demonstrated the feasibility of ML in predicting RPPOA trajectories over a 24-month period ^18,19^. However, the broad clinical translation of these predictive models remains challenging, requiring rigorous validation, interpretability, standardized outcome definitions and careful avoidance of data leakage. Analyzing these longitudinal symptom trajectories requires access to highly standardized, observational data. In this setting, the Osteoarthritis Initiative cohort (OAI) and the Prospective Cohort of Osteoarthritis from A Coruña (PROCOAC) provide a suitable basis for developing a predictive model and testing its performance in an independent European cohort ^20,21^.

Therefore, in this multi-cohort study, we aimed to develop an interpretable multimodal predictive model to identify patients with knee OA at increased risk of RPPOA, and to test its performance in an independent European cohort. This model is intended as a risk-stratification tool to support downstream clinical and research decisions, including closer monitoring, advanced phenotyping, MRI assessment when clinically appropriate and enrichment of clinical trials. By focusing on a clinically meaningful symptomatic outcome and testing the model in an independent cohort, this work aims to support practical risk stratification and future personalized strategies in knee OA.

## METHODS

### Study population

We utilized data from two prospective observational cohorts: the OAI as discovery cohort (initial total N = 3,790), and the PROCOAC as external evaluation cohort (initial total N = 1,511). The OAI includes longitudinal clinical, imaging, and biospecimen data from individuals with or at risk of knee OA (https://nda.nih.gov/oai) ^21^. PROCOAC is a Spanish population-based cohort that tracks OA incidence and progression bi-annually ^20^. Detailed inclusion criteria of this cohort are provided in **Supplementary Methods**. An overview of the complete methodological workflow is illustrated in **Supplementary Figure 1**. All participants provided written informed consent. The study complied with the Declaration of Helsinki and was approved by the Galician Research Ethics Committee (2024/074), utilizing the Rheumatic Diseases Biobank (C.0000424).

### Rapid pain progression definition

The primary outcome, RPPOA, was evaluated across overlapping 24-month observation windows extracted from a 10-year follow-up period (baseline to 120 months). Using the normalized Western Ontario and McMaster (WOMAC) pain score (0-100) ^22^, RPPOA was defined according to IMI-APPROACH definition ^23^ as: (i) an increase _≥_10 points reaching a final score _≥_40; (ii) a rapid increase _≥_20 points reaching a final score _≥_35; or (iii) sustained severe pain (_≥_40 at both timepoints)^18,19^. Overlapping instances were extracted dynamically to accommodate natural clinical variance in visit intervals (20-30 months in PROCOAC).

### Multi-omic integration and preprocessing

OAI genotyping data (dbGAP: phs000955.v1.p1) and PROCOAC samples (Illumina GSA chip) underwent rigorous, identical quality control and imputation protocols. To evaluate previously found genetic signals ^24^, index variants from genome-wide significant loci were extracted via linkage disequilibrium clumping (PLINK 2.0). These variants, alongside mitochondrial ^25^ and quantitative proteomic biomarkers ^26^, were integrated with longitudinal clinical data. Details regarding genomic processing, quality control, and protein quantification are provided in **supplementary methods**.

To ensure genetic homogeneity and account for the specific availability of sequencing and proteomic profiles, the analytical population was restricted to Caucasian participants with valid RPPOA classification.

We consolidated bilateral joint data to retain the “worst-knee” presentation per instance. Features with >50% missingness or near-zero variance, and instances with >40% missing data, were excluded. To prevent data leakage, the cohort was partitioned into training (80%) and testing (20%) sets based on patient IDs, ensuring all longitudinal instances from a single individual remained within the same subset. Missing values were then imputed (linear interpolation for continuous, forward-filling for ordinal, followed by median/mode imputation) using parameters derived strictly from the 80% training set.

### Experimental design and final pipeline assembly

To maximize power while accounting for the varying molecular data availability, we defined two overlapping analytical datasets: a Clinical-Genetic (CG) cohort and a nested Clinical-Genetic-Proteomic (CGP) cohort. To assess the generalizability of the algorithms and build a deployable tool, we employed a dual-pathway design: an “OAI-Complete” pathway (to establish the maximum biological prediction limit) and a “Harmonized” pathway (restricted to features available in PROCOAC).

Model development was encapsulated within scikit-learn pipelines. Five supervised classification algorithms were benchmarked: Random Forest (RF), XGBoost (XGB), Support Vector Machines with Radial Basis Function (SVM-RBF), Multilayer Perceptron Neural Networks (MLP), and Logistic Regression with Elastic Net penalty (EN-LR). To ensure patient independence across folds and prevent data leakage, a 5×5 Repeated Stratified Group K-Fold cross-validation scheme was employed. Models were optimized for the Area Under the Precision-Recall Curve (PR-AUC) to account for class imbalance. Detailed hyperparameter tuning and class imbalance strategies are described in **supplementary methods**.

The algorithm showing optimal balance between cross-validation stability and clinical transparency was selected. As detailed in the Results section, EN-LR emerged as the optimal candidate. To reduce the feature space maintaining key variables, Permutation Feature Importance (PFI) was applied on the training set. Final probabilities were calibrated via nested isotonic regression. Operational decision thresholds were derived from out-of-fold predictions to optimize sensitive screening (F1-score) and high-specificity (F0.5-score).

### Evaluation, model updating and clinical utility

Internal validation was performed on the 20% hold-out test set using cluster bootstrapping (1,000 iterations). Differences between the OAI-Complete and Harmonized pathways were assessed via cluster-paired permutation testing. Feature importance and the directional impact of predictors were quantified using SHapley Additive exPlanations (SHAP) values.

For external evaluation of the model, the frozen Harmonized pipeline was deployed on PROCOAC. Recognizing the baseline prevalence shift between cohorts (OAI ∼10% vs. PROCOAC ∼28%), we applied post-hoc logistic recalibration (intercept and slope updating). Calibration performance pre- and post-updating was assessed using the Observed-to-Expected (O:E) ratio and the Brier score. Clinical utility was assessed via Decision Curve Analysis (DCA) to quantify Net Benefit against default strategies. Finally, the internally derived thresholds were applied to the externally recalibrated probabilities to stratify individuals into three distinct risk categories (low, intermediate and high) to guide clinical action. The overall study design, model development, external evaluation, and reporting adhered strictly to the TRIPOD+AI guidelines ^27^ **(Supplementary Table 2)**.

## RESULTS

### Study population

The analytical population in the discovery cohort (OAI) comprised 2,934 Caucasian individuals (14,488 longitudinal instances) for the primary CG analysis, and 1,130 individuals (6,050 instances) for the nested CGP analysis. The external evaluation cohort (PROCOAC) included 582 individuals (1,609 instances) and 299 individuals (845 instances) for the CG and CGP cohorts, respectively (**Table 1**).

**Table 1.** Baseline characteristics of the discovery (OAI) and external evaluation (PROCOAC) cohorts. *.

| Baseline Characteristic | OAI CG cohort<br>(N = 2,934) | OAI CGP cohort<br>(N = 1,130) | PROCOAC CG cohort<br>(N = 582) | PROCOAC CGP cohort<br>(N = 299) |
| --- | --- | --- | --- | --- |
| **Demographics (at baseline)** |  |  |  |  |
| Age (years), mean (SD) | 61.8 (9.2) | 60.1 (8.9) | 63.7 (8.4) | 64.8 (8.6) |
| Female sex, N (%) | 1,627 (55.5%) | 589 (52.1%) | 459 (78.9%) | 242 (80.9%) |
| Body mass index (kg/m<sup>2</sup>), mean (SD) | 27.5 (0.8) | 27.3 (0.7) | 29.3 (5.0) | 30.0 (5.2) |
| **Clinical variables (at baseline)** |  |  |  |  |
| Normalized WOMAC pain score (0-100), median [IQR] | 5 [0 - 20] | 5 [0 - 20] | 4 [0 - 7] | 5 [1 - 8] |
| KL grade (0-4) | 2 [0 - 2] | 1 [0 - 2] | 2 [1 - 2] | 2 [1 - 3] |
| **Longitudinal follow-up & outcome** |  |  |  |  |
| Total longitudinal instances, N | 14,488 | 6,050 | 1,609 | 845 |
| Instances per patient, mean (SD) | 4.9 (1.3) | 5.3 (1.0) | 2.8 (1.3) | 2.8 (1.3) |
| Rapid pain progression, N (%)<sup>†</sup> | 1,473 (10.2%) | 639 (10.6%) | 448 (27.8%) | 255 (30.2%) |

\* Baseline values refer to the first available visit for each unique patient. CG denotes Clinical-Genetic; CGP, Clinical-Genetic-Proteomic; IQR, Interquartile Range; KL, Kellgren-Lawrence; OAI, Osteoarthritis Initiative; PROCOAC, Prospective Cohort of Osteoarthritis from A Coruña; SD, Standard Deviation; and WOMAC, Western Ontario and McMaster Universities Osteoarthritis Index.
† Rapid pain progression is calculated over the total number of longitudinal 24-month instances rather than unique patients.

The overall baseline prevalence of RPPOA was approximately 10% across both the CG (10.2%) and the CGP (10.6%) cohorts in the OAI. In contrast, PROCOAC exhibited a distinct epidemiological profile characterized by older population, with higher Body Mass Index (BMI), a greater proportion of females, and a notably higher RPPOA prevalence (27.8% in CG cohort).

### Algorithm benchmarking and feature selection

During initial cross-validation, EN-LR and RF demonstrated optimal scores and stability **(Supplementary Figure 2).** EN-LR was selected as the optimal algorithm due to its added simplicity and interpretability. To transparently report the baseline capabilities of the unpruned algorithms, their initial performance was evaluated on the independent hold-out test set (**Figure 1**).

**Figure 1.**
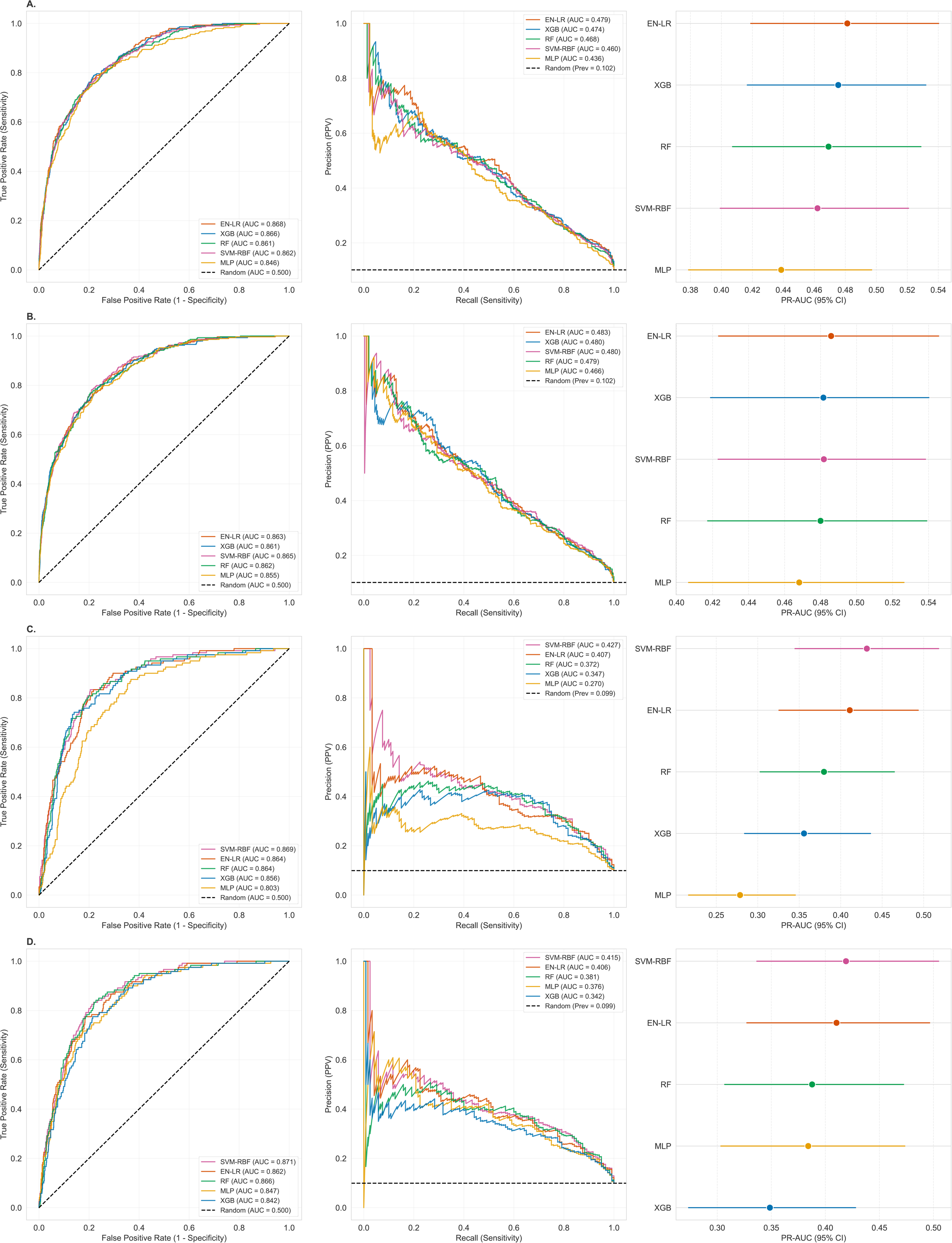
Machine learning performance benchmarking across dataset configurations in Hold-out Test set. A) CG Cohort OAI-Complete; B) CG Cohort harmonized; C) CGP Cohort OAI-Complete; D) CGP Cohort harmonized. Each row displays the baseline predictive performance of five distinct machine learning algorithms (EN-LR, XGB, RF, SVM-RBF, and MLP) on the hold-out test set prior to feature selection and hyperparameter fine-tuning. The left column presents Receiver Operating Characteristic (ROC) curves with their respective Area Under the Curve (AUC) scores. The center column shows Precision-Recall (PR) curves, which are more informative given the outcome prevalence (represented by the black dashed line). The right column displays a forest plot of the PR-AUC point estimates and their 95% Confidence Intervals (calculated via bootstrapping), providing a statistical comparison of algorithm performance for each cohort variation. CG denotes Clinical-Genetic; CGP, Clinical-Genetic-Proteomic; OAI, Osteoarthritis Initiative; EN-LR, Elastic-Net Logistic Regression; RF, Random Forest; XGB, eXtreme Gradient Boosting; MLP, Multilayer Perceptron; and SVM-RBF, Support Vector Machine with Radial Basis Function kernel.

As a result of applying PFI, the OAI-complete CG feature space was reduced from 354 to 20 final variables, while the harmonized CG feature space was reduced from 159 to 19 critical predictors. Within the nested CGP cohort, PFI systematically discarded all serum proteomic variables. Given this lack of incremental predictive value from baseline proteomics and the smaller sample of individuals, subsequential evaluation was restricted to the primary CG pipelines. Following final hyperparameter re-optimization **(Supplementary Table 3)**, the harmonized signature was driven by baseline radiographic severity (Kellgren-Lawrence grade), mechanical joint pain, WOMAC stiffness, BMI, and two genetic variants (rs73631790, rs9912678) (**Figure 2, Supplementary Figure 3**).

**Figure 2.**
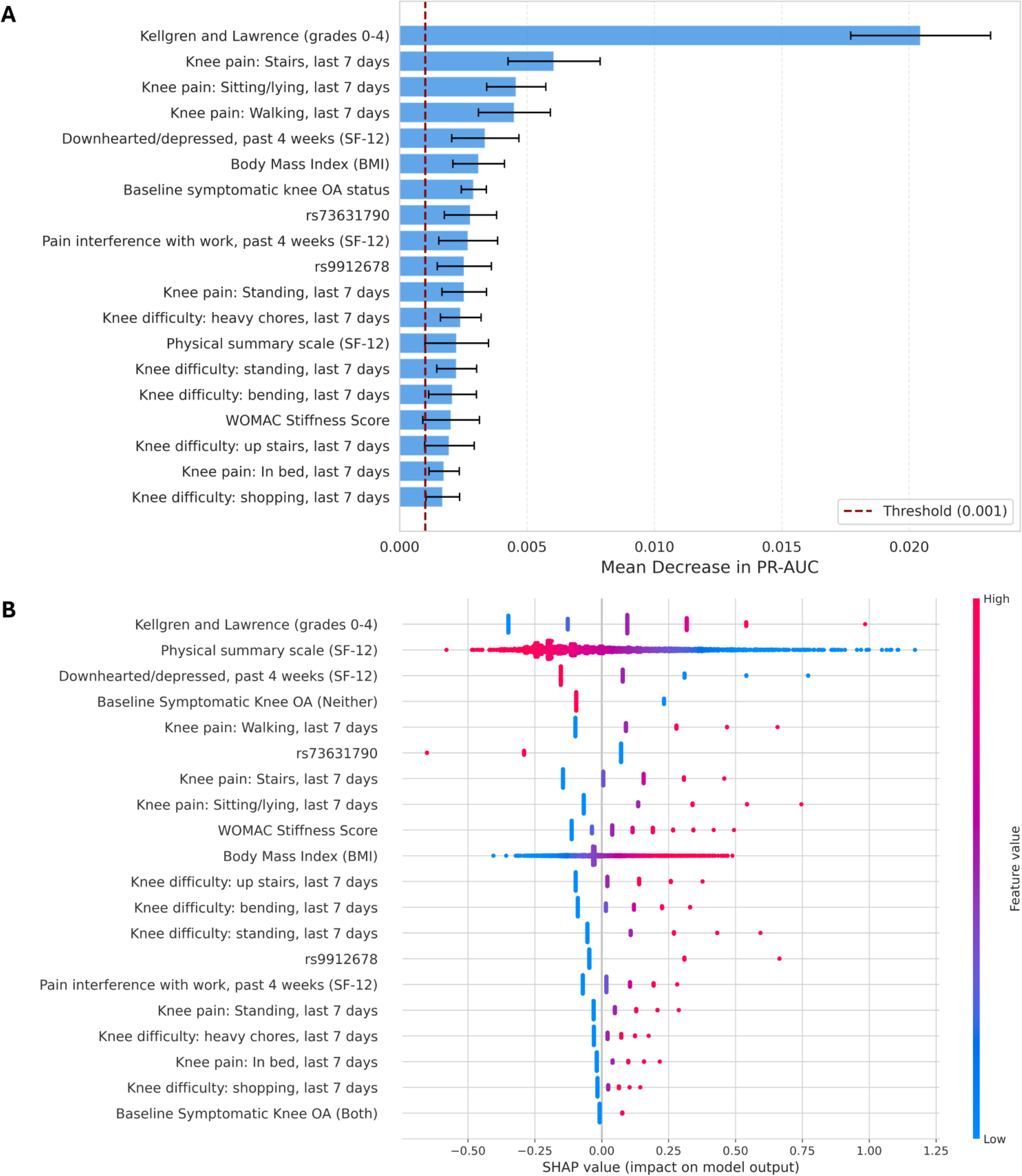
Feature importance and Global interpretability of the harmonized predictive model in the Clinical Genetic (CG) Cohort. (A) Permutation feature importance ranking for the primary GC Cohort. Variables are sorted by their mean impact on the model’s predictive performance (PR-AUC decrement) when permuted. Error bars represent the 95% confidence intervals across repeated permutations. (B) SHapley Additive exPlanations (SHAP) summary dot plot evaluating the directionality and magnitude of each feature’s contribution to the prediction of rapid pain progression. Each dot represents a single patient’s hold-out test evaluation. Colour indicates the actual feature value (red = high, blue = low), and the position on the x-axis indicates the SHAP value (impact on the probability of rapid pain progression). OA denotes Osteoarthritis; SF-12, 12-Item Short Form Health Survey; and WOMAC, Western Ontario and McMaster Universities Osteoarthritis Index.

### Internal Validation and Cost of Generalizability

On the internal hold-out test set, restricting the model to externally available features did not penalize performance. The Harmonized pipeline achieved a PR-AUC of 0.478 (95% CI: 0.399-0.552) and a ROC-AUC of 0.861 (95% CI: 0.831-0.886), establishing statistical non-inferiority compared to the unrestricted OAI-Complete pathway (PR-AUC 0.447, 95% CI: 0.373-0.519; cluster-paired permutation p = 0.060) (**Table 2**).

**Table 2.**
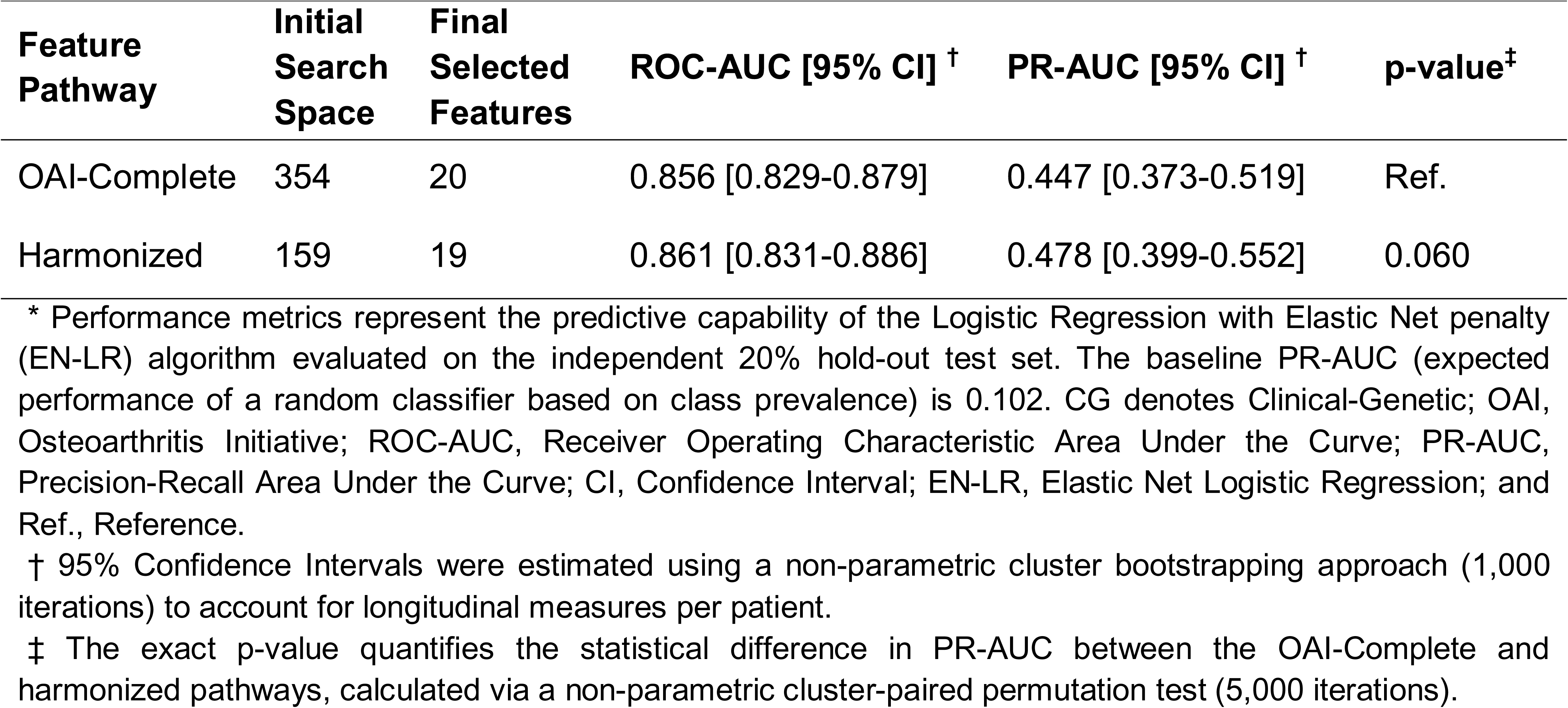
Internal validation and discriminative performance of the predictive pipelines in the Clinical-Genetic (CG) Cohort. *.

| **Feature Pathway** | **Initial Search Space** | **Final Selected Features** | **ROC-AUC [95% CI] <sup>†</sup>** | **PR-AUC [95% CI] <sup>†</sup>** | **p-value<sup>‡</sup>** |
| --- | --- | --- | --- | --- | --- |
| OAI-Complete | 354 | 20 | 0.856 [0.829-0.879] | 0.447 [0.373-0.519] | Ref. |
| Harmonized | 159 | 19 | 0.861 [0.831-0.886] | 0.478 [0.399-0.552] | 0.060 |

\* Performance metrics represent the predictive capability of the Logistic Regression with Elastic Net penalty (EN-LR) algorithm evaluated on the independent 20% hold-out test set. The baseline PR-AUC (expected performance of a random classifier based on class prevalence) is 0.102. CG denotes Clinical-Genetic; OAI, Osteoarthritis Initiative; ROC-AUC, Receiver Operating Characteristic Area Under the Curve; PR-AUC, Precision-Recall Area Under the Curve; CI, Confidence Interval; EN-LR, Elastic Net Logistic Regression; and Ref., Reference.
<sup>†</sup> 95% Confidence Intervals were estimated using a non-parametric cluster bootstrapping approach (1,000 iterations) to account for longitudinal measures per patient.
<sup>‡</sup> The exact p-value quantifies the statistical difference in PR-AUC between the OAI-Complete and harmonized pathways, calculated via a non-parametric cluster-paired permutation test (5,000 iterations).

### External Evaluation and Model Updating

When deployed on the independent PROCOAC cohort, the frozen harmonized model demonstrated robust cross-continental transportability, maintaining high discrimination (ROC-AUC 0.744, 95% CI: 0.714-0.772). Driven by the higher baseline prevalence in the PROCOAC cohort, the model’s Precision-Recall performance improved to a PR-AUC of 0.519 (95% CI: 0.458–0.581) (**Table 3**). However, direct geographical translation induced calibration drift, causing the raw model to underestimate PROCOAC risk (O:E ratio 1.348). Following logistic recalibration, probability outputs were restored (O:E ratio 1.000, Scaled Brier score 0.118; **Supplementary Table 4**) without altering the model’s underlying discriminative ranking (**Figure 3A**).

**Table 3.**
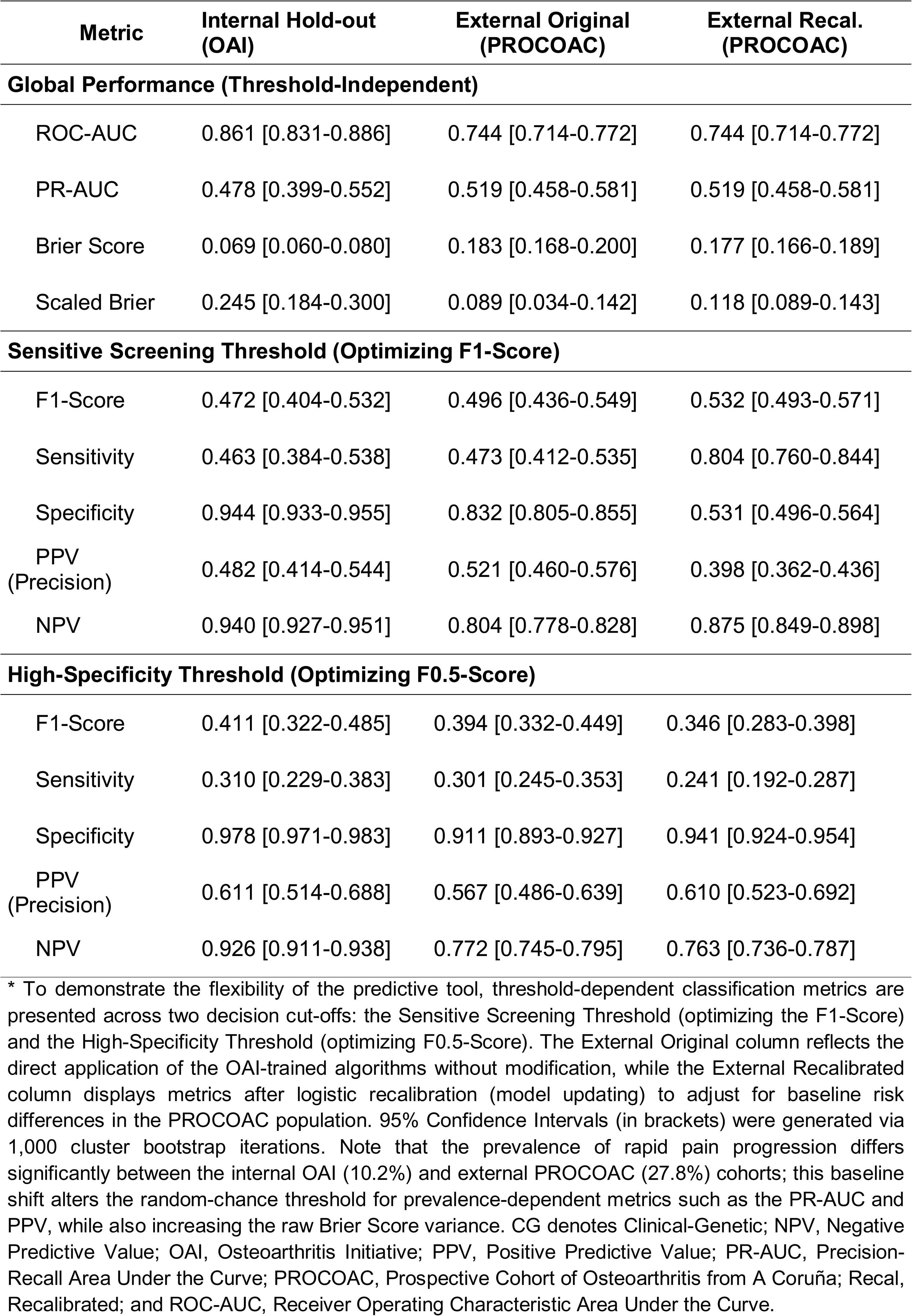
Internal versus external evaluation cohort and model updating metrics of the optimal harmonized pipeline for the CG Cohort evaluated at distinct operational thresholds. *.

**Figure 3.**
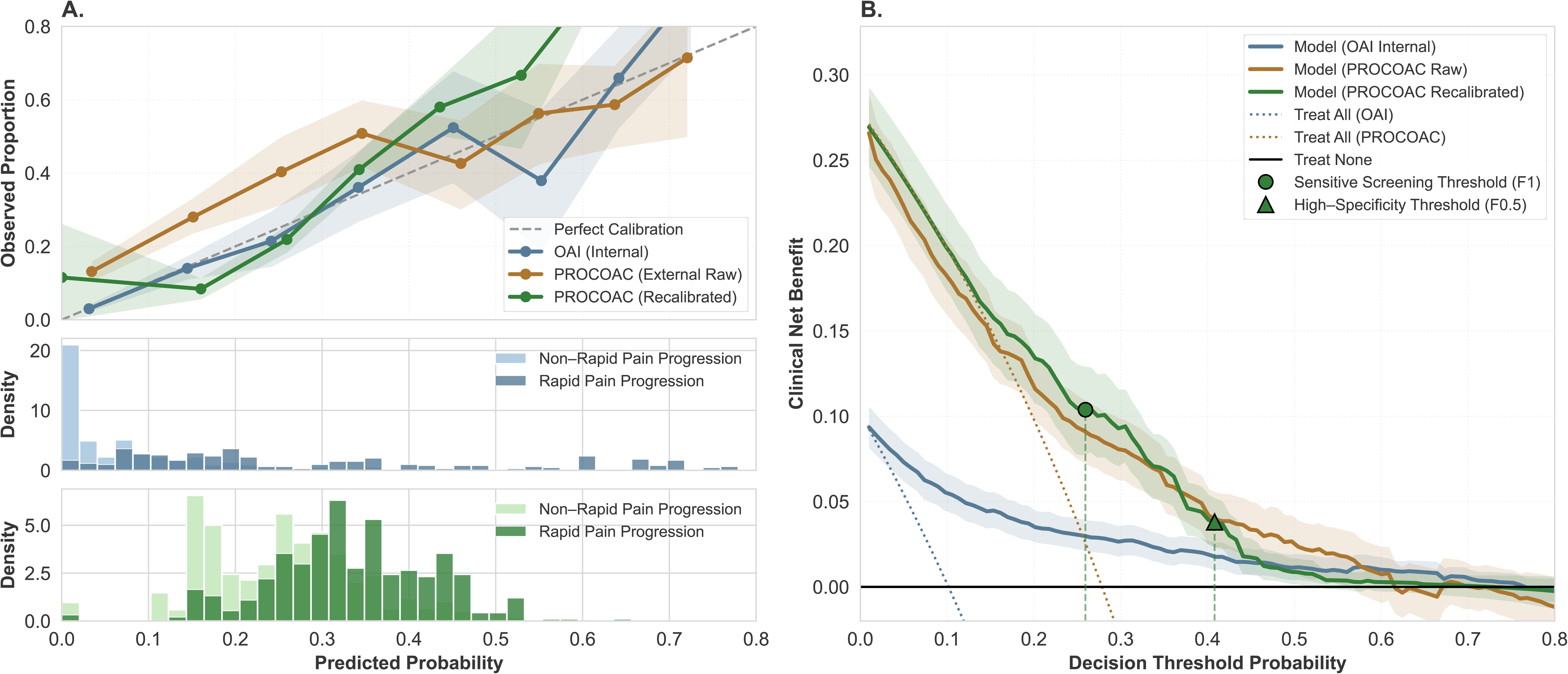
Clinical utility, calibration, and probability distributions of the harmonized predictive pipeline in the Clinical-Genetic (CG) Cohort. (A) Calibration plots comparing the internal hold-out validation (OAI, blue), external raw application (PROCOAC, orange), and external recalibrated model (PROCOAC, green). Observed proportions with 95% confidence intervals are plotted against predicted probabilities. The bottom sub-panels display the probability density distributions for the internal OAI (blue) and recalibrated PROCOAC (green) cohorts to account for class imbalance and differing baseline prevalences (∼10% vs. ∼28%). (B) Decision Curve Analysis (DCA) illustrating the Clinical Net Benefit of utilizing the harmonized model (coloured lines) across decision threshold probabilities, compared against alternative default strategies: assuming all patients will experience rapid progression (“Treat All”, dotted lines) or assuming no patients will experience it (“Treat None”, horizontal black line). Shaded areas represent 95% confidence intervals derived from 1,000 bootstrap iterations. The projected symbols on the recalibrated curve indicate the operational thresholds mapped for sensitive screening (circle at 25.9%) and high-specificity prioritization (triangle at 40.8%).

### Clinical utility and risk stratification

DCA confirmed that the recalibrated model provided substantial positive Net Benefit across a clinically relevant risk range (10% to 50%), consistently outperforming default “treat-all” and “treat-none” default strategies (**Figure 3B**). Evaluating the previously defined operational thresholds (**Supplementary Table 5**), the recalibrated model achieved a Net Benefit of approximately 0.10 at the sensitive screening threshold (25.9%), with the lower bound of its 95% confidence interval remaining strictly above the default approaches. At the stricter high-specificity threshold (40.8%), the model maintained a positive Net Benefit of approximately 0.04. The uncalibrated model yielded consistently lower Net Benefit across all thresholds, supporting the need for recalibration before applying predicted probabilities in the external cohort.

Application of the predefined risk-stratification framework to the recalibrated external probabilities defined three clinically interpretable strata. Patients below the sensitive screening threshold (<0.26) formed a low-risk stratum characterized by an NPV of 0.875 (95% CI: 0.849–0.898), identifying a subgroup in whom 24-month RPPOA was relatively unlikely. Patients above the high-specificity threshold (_≥_0.40) formed a high-risk stratum, with specificity 0.941 (95% CI: 0.924–0.954) and PPV 0.610 (95% CI: 0.523–0.692), identifying a subgroup enriched for RPPOA and potentially suitable for downstream clinical or research prioritization. Patients between these cut-offs (0.26–<0.40) constituted an intermediate-risk stratum, reflecting a zone of predictive uncertainty where treatment optimization, closer monitoring, and short-term reassessment may be appropriate (**Supplementary Table 6**).

## DISCUSSION

In this multi-cohort study, we developed an interpretable ML pipeline to predict RPPOA over 24-month windows and independently tested its performance in PROCOAC. By reducing hundreds of multimodal variables, we identified a robust 19-variable Clinical-Genetic signature comprising 17 standard clinical and demographics assessments (including Kellgren-Lawrence grade, localized knee pain, and BMI) alongside two genetic variants (rs73631790 and rs9912678). This penalized logistic regression approach maintained good discrimination in the OAI hold-out set and in the independent PROCOAC cohort, while recalibration improved the agreement between predicted and observed risk. To ensure real-world utility, we translated predictive probabilities into a practical risk stratification. By setting specific thresholds for primary care screening and high-specificity prioritization, we provide an actionable tool to guide clinical decisions and optimize healthcare resources in OA management.

Recent machine learning advancements predict OA progression; however, our framework prioritizes clinical accessibility and phenotypic isolation. Contemporary deep learning frequently relies on predicting structural deterioration using complex MRI features or multimodal radiographs ^28–30^. While offering mechanistic insights, their dependence on advanced imaging makes routine primary care screening economically and logistically unfeasible. Furthermore, existing models often conflate structural decay with symptom worsening. Given the well-documented discordance between radiographic joint space narrowing and patient-reported pain ^31,32^, our work isolates RPPOA. Focusing exclusively on this highly unstable symptomatic trajectory directly addresses the primary driver of patient disability, providing a more actionable endpoint for patient management and trial enrichment ^14^.

A critical methodological strength was algorithmic transparency. Although most tested algorithms (e.g., Random Forest, SVM-RBF) achieved comparable discriminative power, their “black-box” nature, similar to dual-classifiers ^19^ or heavily weighted AutoML ensembles ^18^, hinders physician trust and clinical deployment. By selecting EN-LR leaning toward a Ridge-like (L2) penalty, we effectively managed multicollinearity and reduced dimensionality relying on PFI, while obtaining a transparent, parsimonious signature easily interpretable by healthcare professionals. Notably, the OAI-Complete and harmonized models revealed practically identical performance, demonstrating that excluding cohort-specific variables did not sacrifice predictive power but rather acted as an effective noise-reduction mechanism.

The retention of genetic variants proximal to the *DUXA* (rs73631790) and *RAI1* (rs9912678) genes ^24^ over competing clinical predictors suggests they harbor independent prognostic value for RPPOA. *RAI1* is implicated in nociceptive and circadian dysregulation, directly influencing pain thresholds ^33^, while the *DUX* family, which includes *DUXA,* is linked to oxidative stress and mitochondrial dysfunction in musculoskeletal tissues ^34^. Collectively, this indicates that although macroscopic damage and biomechanical overload drive imminent pain exacerbation^3^, underlying genetic vulnerabilities to sensory and redox imbalances act as measurable mechanistic mediators.

Our study also addresses geographic portability, a critical translational bottleneck in medical artificial intelligence (AI) ^35^. As previously reviewed ^36^, few OA predictive algorithms achieve true external validation, and those typically target endpoints like structural deterioration or joint replacement ^37–41^. In contrast, we provide cross-continental evaluation by deploying our OAI-trained algorithm on an independent European cohort (PROCOAC). In doing so, we directly confronted the phenomenon of calibration drift induced by epidemiological shift (RPPOA prevalence: ∼10% US vs. ∼30% Spain). Adhering to TRIPOD+AI guidelines ^27^, post-hoc logistic recalibration of the baseline risk successfully rescued the miscalibrated predictions. This demonstrates that algorithmic transportability does not require costly retraining from scratch; transparent models may be transported across cohorts when appropriate recalibration is performed, although prospective implementation studies are still required before routine clinical use.

For clinical implementation, we propose categorizing the recalibrated probabilities into three pragmatic risk strata **(Supplementary Table 6)**. Patients with an estimated 24-month risk below 0.26 constitute the low-risk group in whom standard knee OA management and routine follow-up may be appropriate, provided there are no atypical features or other clinical indications for advanced assessment. Patients between 0.26 and 0.40 represent an intermediate risk group, in whom treatment optimization, closer monitoring and short-term reassessment may be warranted. Finally, individuals with predicted risk _≥_0.40 constitute a high-risk group enriched for RPPOA, for whom advanced phenotyping, MRI assessment when clinically appropriate or referral to clinical trials could be prioritized.

Several limitations must be acknowledged. First, restricting the cohort to Caucasian individuals to prevent genetic stratification leaves the pipeline’s transportability to diverse ethnicities untested ^42^. Second, measuring molecular biomarkers exclusively at baseline precluded capturing protein expression changes over time, likely reducing the algorithm’s ability to use dynamic proteomic signals to predict rapid symptom changes ^43^. Third, the utilized genomic features derive from a GWAS with a relatively modest sample size ^24,44^. Fourth, while recalibration optimized the external screening profile, it mathematically increased the false-positive rate at lower decision thresholds. Finally, real-time prospective clinical deployments remain necessary to confirm the tool’s real-world utility ^45,46^.

Despite these limitations, this study presents several major strengths. To our knowledge, this is one of the first OA machine-learning predictive models integrating clinical and molecular parameters to undergo rigorous, cross-continental external testing in an independent prospective European cohort (PROCOAC). Our comprehensive methodology-encompassing explainable AI, formal DCA, adherence to recalibration protocols, and actionable operational thresholds bridges the gap between mathematical performance and real clinical utility, providing a reliable foundation for future personalized medicine tools. In conclusion, this interpretable predictive pipeline identifies patients at risk of RPPOA within 24-month windows using a parsimonious MRI-free signature driven by accessible clinical variables and targeted genetic information. By translating recalibrated probabilities into three clinically interpretable strata, the model provides a practical framework for risk-adapted management, closer follow-up and selection of patients for advanced phenotyping or clinical trials. Prospective implementation studies are still required before routine clinical use, but these findings support the potential value of externally tested, interpretable prediction models for personalized care in knee OA.

## Data Availability

All data produced in the present study are available upon reasonable request to the authors

## Funding

Supported by funding from the Instituto de Salud Carlos III through projects RD24/0007/0026, PMP22/00101, PMPTA22/00115, PI22/01155, PI23/00818, and PI23/00913, and co-funded by the European Union. This work was also funded by grants IN607A2025/11 from the Axencia Galega de Innovación-Xunta de Galicia. J.V.-G. was supported by grant IN606A 2022/048 from the Xunta de Galicia. L.M.-S. was supported by an i-PFIS predoctoral fellowship from the Instituto de Salud Carlos III (IFI24/00042). Additional funding was provided by Pfizer and Eli Lilly and Company through the 3rd Global Awards for Advancing Chronic Pain Research (ADVANCE, grant ID#64122119).

## Conflict of interests

We declare no competing interests.

## Acknowledgements

The authors are indebted to the study coordinators, research staff, and participants of both the Prospective Cohort of Osteoarthritis from A Coruña (PROCOAC) and the Osteoarthritis Initiative (OAI). The OAI is a public-private partnership comprised of five contracts (N01-AR-2-2258; N01-AR-2-2259; N01-AR-2-2260; N01-AR-2-2261; N01-AR-2-2262) funded by the National Institutes of Health (NIH), a branch of the Department of Health and Human Services and conducted by the OAI Study Investigators. Private funding partners include Pfizer, Inc.; Novartis Pharmaceuticals Corporation; Merck Research Laboratories; and GlaxoSmithKline. Private sector funding for the OAI is managed by the Foundation for the National Institutes of Health. This manuscript was prepared using an OAI public use data set and does not necessarily reflect the opinions or views of the OAI investigators, the NIH, or the private funding partners.

Additionally, during the preparation of this manuscript, the authors utilized ChatGPT to assist with the refinement of R and Python scripts, as well as to improve the English language and structural flow of the text. Following the use of this tool, the authors rigorously reviewed and edited all material and take full responsibility for the final content and integrity of the publication.

## Notes

### Competing Interest Statement

The authors have declared no competing interest.

### Author Declarations

The study complied with the Declaration of Helsinki and was approved by the Galician Research Ethics Committee (2024/074), utilizing the Rheumatic Diseases Biobank (C.0000424)

