## Supplemental Tables for "Risk stratification for the rapid pain progression phenotype in knee osteoarthritis using interpretable multimodal machine learning: Development in the Osteoarthritis Initiative and external evaluation in the Prospective Cohort of Osteoarthritis from A Coruña"

**Supplementary tables**

| **Supplementary Table 1. Hyperparameter search space defined for the algorithm benchmarking phase.** | | | | |
| --- | --- | --- | --- | --- |
| **Algorithm** | **Hyperparameter** | **Description** | **Evaluated Search Space** | **Fixed Parameters** |
| Random Forest (RF) | n_estimators | Number of trees in the forest | 200, 300, 500, 800 | class_weight: 'balanced' |
|  | max_depth | Maximum depth of the trees | 3, 5, 7, 10 |  |
|  | min_samples_leaf | Minimum samples required at a leaf node | 10, 20, 30 |  |
|  | max_samples | Fraction of samples used for bagging | 0.6, 0.8, None (1.0) |  |
|  | max_features | Number of features to consider per split | 'sqrt', 'log2', 0.3, 0.5 |  |
|  | criterion | Split quality criterion | 'gini', 'entropy' |  |
| Elastic Net Logistic Regression (EN-LR) | C | Inverse of regularization strength | 20 log-spaced values from 0.001 to 100 | penalty: 'elasticnet';  solver: 'saga';  class_weight: 'balanced';  max_iter: 50,000 |
|  | l1_ratio | Elastic Net mixing parameter | 11 linear-spaced values from 0.0 to 1.0 |  |
| XGBoost (XGB) | n_estimators | Number of boosting rounds | 100, 200, 300, 500 | objective: 'binary: logistic';  tree_method: 'hist' |
|  | learning_rate | Step size shrinkage used in update | 0.01, 0.05, 0.1 |  |
|  | max_depth | Maximum tree depth for base learners | 3, 4, 5, 6 |  |
|  | subsample | Subsample ratio of the training instances | 0.6, 0.8, 1.0 |  |
|  | colsample_bytree | Subsample ratio of columns per tree | 0.6, 0.8, 1.0 |  |
|  | min_child_weight | Minimum sum of instance weight in a child | 1, 5, 10 |  |
|  | reg_alpha | L1 regularization term on weights | 0, 0.1, 1, 5 |  |
|  | reg_lambda | L2 regularization term on weights | 0, 1, 5, 10 |  |
|  | scale_pos_weight | Balancing of positive and negative weights | 5, 8, 9, 12 |  |
| Support Vector Machine (SVM-RBF) | C | Regularization parameter | 0.1, 0.5, 1, 3, 5 | kernel: 'rbf';  probability: True |
|  | gamma | Kernel coefficient | 'scale', 0.1, 0.001 |  |
|  | class_weight | Penalty weights per class | 'balanced', {0:1, 1:5}, {0:1, 1:9} |  |
| Multilayer Perceptron (MLP) | hidden_layer_sizes | Number of neurons per hidden layer | (100,), (50, 25) | solver: 'adam';  learning_rate: 'adaptive';  max_iter: 1,000;  early_stopping: True |
|  | activation | Activation function for hidden layers | 'relu', 'tanh' |  |
|  | alpha | L2 regularization penalty | 0.1, 1, 2, 5 |  |
|  | learning_rate_init | Initial learning rate | 0.001, 0.01 |  |
| The hyperparameter optimization during the benchmarking phase was conducted using a RandomizedSearchCV strategy with 50 iterations per algorithm, evaluated via a 5x5 Repeated Stratified Group K-Fold Cross-Validation scheme to maximize the Precision-Recall Area Under the Curve (PR-AUC). For EN-LR, the C parameter grid was generated using NumPy's logspace function, and the l1_ratio was generated using linspace. | | | | |

| **Supplementary Table 2. TRIPOD+AI checklist for the reporting of prediction model studies** | | | | |
| --- | --- | --- | --- | --- |
| **Section/topic** | **Item** | **D/E*** | **Checklist item** | **Page/Section** |
| **Title** | | | | |
| Title | 1 | D;E | Identify the study as developing or evaluating the performance of a multivariable prediction model, the target population, and the outcome to be predicted | 1 |
| **Abstract** | | | | |
| Abstract | 2 | D;E | Provide a structured summary encompassing the study identification, background, objectives, methods (data, eligibility, outcome, modelling, and metrics), results (sample size, predictors, and performance estimates), interpretation, and registration details | 2 |
| **Introduction** | | | | |
| Background | 3a | D;E | Explain the healthcare context (including whether diagnostic or prognostic) and rationale for developing or evaluating the prediction model, including references to existing models | 4-5 |
|  | 3b | D;E | Describe the target population and the intended purpose of the prediction model in the context of the care pathway, including its intended users (eg, healthcare professionals, patients, public) | 5 |
|  | 3c | D;E | Describe any known health inequalities between sociodemographic groups | 15 |
| Objectives | 4 | D;E | Specify the study objectives, including whether the study describes the development or validation of a prediction model (or both) | 5 |
| **Methods** | | | | |
| Data | 5a | D;E | Describe the sources of data separately for the development and evaluation datasets (eg, randomised trial, cohort, routine care or registry data), the rationale for using these data, and representativeness of the data | 6 & supplementary methods 3 |
|  | 5b | D;E | Specify the dates of the collected participant data, including start and end of participant accrual; and, if applicable, end of follow-up | Supplementary methods 3 |
| Participants | 6a | D;E | Specify key elements of the study setting (eg, primary care, secondary care, general population) including the number and location of centres | 6 & 7 |
|  | 6b | D;E | Describe the eligibility criteria for study participants | Supplementary methods 3 |
|  | 6c | D;E | Give details of any treatments received, and how they were handled during model development or evaluation, if relevant | N/A (Observational cohort, standard care) |
| Data Preparation | 7 | D;E | Describe any data pre-processing and quality checking, including whether this was similar across relevant sociodemographic groups | 8 & supplementary methods 1/3 |
| Outcome | 8a | D;E | Clearly define the outcome that is being predicted and the time horizon, including how and when assessed, the rationale for choosing this outcome, and whether the method of outcome assessment is consistent across sociodemographic groups | 6 |
|  | 8b | D;E | If outcome assessment requires subjective interpretation, describe the qualifications and demographic characteristics of the outcome assessors | N/A (standardized self-reported WOMAC questionnaire) |
|  | 8c | D;E | Report any actions to blind assessment of the outcome to be predicted | N/A |
| Predictors | 9a | D | Describe the choice of initial predictors (eg, literature, previous models, all available predictors) and any pre-selection of predictors before model building | 7 & supplementary methods 3 |
|  | 9b | D;E | Clearly define all predictors, including how and when they were measured (and any actions to blind assessment of predictors for the outcome and other predictors) | Supplementary methods 1-S3 |
|  | 9c | D;E | If predictor measurement requires subjective interpretation, describe the qualifications and demographic characteristics of the predictor assessors | Supplementary methods 3 (KL grading) |
| Sample Size | 10 | D;E | Explain how the study size was arrived at (separately for development and evaluation), and justify that the study size was sufficient to answer the research question. Include details of any sample size calculation | 6 & 7 (All eligible cohort data used) |
| Missing data | 11 | D;E | Describe how missing data were handled. Provide reasons for omitting any data | 7 & supplementary methods 3 |
| Analytical methods | 12a | D | Describe how the data were used (eg, for development and evaluation of model performance) in the analysis, including whether the data were partitioned, considering any sample size requirements | 7 & supplementary methods 3 |
|  | 12b | D | Depending on the type of model, describe how predictors were handled in the analyses (functional form, rescaling, transformation, or any standardisation) | Supplementary methods 5 |
|  | 12c | D | Specify the type of model, rationale†, all model building steps, including any hyperparameter tuning, and method for internal validation | 8 & supplementary methods 5 |
|  | 12d | D;E | Describe if and how any heterogeneity in estimates of model parameter values and model performance was handled and quantified across clusters (eg, hospitals, countries). | Supplementary methods 6 (Cluster bootstrapping) |
|  | 12e | D;E | Specify all measures and plots used (and their rationale) to evaluate model performance (eg, discrimination, calibration, clinical utility) and, if relevant, to compare multiple models | 8-9 & supplementary methods 6 |
|  | 12f | E | Describe any model updating (eg, recalibration) arising from the model evaluation, either overall or for particular sociodemographic groups or settings | 8-9 & supplementary methods 7 |
|  | 12g | E | For model evaluation, describe how the model predictions were calculated (eg, formula, code, object, application programming interface) | Supplementary methods 5 |
| Class Imbalance | 13 | D;E | If class imbalance methods were used, state why and how this was done, and any subsequent methods to recalibrate the model or the model predictions | Supplementary methods 5 |
| Fairness | 14 | D;E | Describe any approaches that were used to address model fairness and their rationale | 15 (Limitations: restricting to Caucasian population) |
| Model output | 15 | D | Specify the output of the prediction model (eg, probabilities, classification). Provide details and rationale for any classification and how the thresholds were identified | 8 |
| Training vs. evaluation | 16 | D;E | Identify any differences between the development and evaluation data in healthcare setting, eligibility criteria, outcome, and predictors | 8-9 & table 1 |
| Ethical approval | 17 | D;E | Name the institutional research board or ethics committee that approved the study and describe the participant informed consent or the ethics committee waiver of informed consent | 6 & 16 |
| **Open Science** | | | | |
| Funding | 18a | D;E | Give the source of funding and the role of the funders for the present study | 1 & 16 |
| Conflicts of interest | 18b | D;E | Declare any conflicts of interest and financial disclosures for all authors | 2 |
| Protocol | 18c | D;E | Indicate where the study protocol can be accessed or state that a protocol was not prepared | N/A (Retrospective observational analysis) |
| Registration | 18d | D;E | Provide registration information for the study, including register name and registration number, or state that the study was not registered | N/A (Retrospective observational analysis) |
| Data sharing | 18e | D;E | Provide details of the availability of the study data | 16 |
| Code sharing | 18f | D;E | Provide details of the availability of the analytical code§ | N/A (Code available upon reasonable request) |
| **Patient and public involvement** | | | | |
| Patient and public involvement | 19 | D;E | Provide details of any patient and public involvement during the design, conduct, reporting, interpretation, or dissemination of the study or state no involvement | N/A |
| **Result** | | | | |
| Participants | 20a | D;E | Describe the flow of participants through the study, including the number of participants with and without the outcome and, if applicable, a summary of the follow-up time. A diagram may be helpful | 9 & table 1 |
|  | 20b | D;E | Report the characteristics overall and, where applicable, for each data source or setting, including the key dates, key predictors (including demographics), treatments received, sample size, number of outcome events, follow-up time, and amount of missing data. A table may be helpful. Report any differences across key demographic groups | 9 & table 1 |
|  | 20c | E | For model evaluation, show a comparison with the development data of the distribution of important predictors (demographics, predictors, and outcome) | Table 1 |
| Model development | 21 | D;E | Specify the number of participants and outcome events in each analysis (eg, for model development, hyperparameter tuning, model evaluation) | 9 & table 1 |
| Model specification | 22 | D | Provide details of the full prediction model (eg, formula, code, object, application programming interface) to allow predictions in new individuals and to enable third party evaluation and implementation, including any restrictions to access or reuse (eg, freely available, proprietary)¶ | Supplementary methods 5, supplementary tables 1, 3 & 5 |
| Model performance | 23a | D;E | Report model performance estimates with confidence intervals, including for any key subgroups (eg, sociodemographic). Consider plots to aid presentation | 10-11 & tables 2-3 |
|  | 23b | D;E | If examined, report results of any heterogeneity in model performance across clusters. | N/A |
| Model updating | 24 | E | Report the results from any model updating, including the updated model and subsequent performance | 11, figure 4, table 3 & supplementary table 4 |
| **Discussion** | | | | |
| Interpretation | 25 | D;E | Give an overall interpretation of the main results, including issues of fairness in the context of the objectives and previous studies | 12-15 |
| Limitations | 26 | D;E | Discuss any limitations of the study (such as a non-representative sample, sample size, overfitting, missing data) and their effects on any biases, statistical uncertainty, and generalisability | 15 |
| Usability | 27a | D | Describe how poor quality or unavailable input data (eg, predictor values) should be assessed and handled when implementing the prediction model | Supplementary methods 3 |
|  | 27b | D | Specify whether users will be required to interact in the handling of the input data or use of the model, and what level of expertise is required of users | 14-15 & table 4 |
|  | 27c | D;E | Discuss any next steps for future research, with a specific view to applicability and generalisability of the model | 15 |
| TRIPOD denotes Transparent Reporting of a multivariable prediction model for Individual Prognosis Or Diagnosis; AI, artificial intelligence; *D, items relevant only to the development of a prediction model; E, items relating solely to the evaluation of a prediction model; D;E, items applicable to both the development and evaluation of a prediction model; WOMAC, Western Ontario and McMaster Universities Osteoarthritis Index; and KL, Kellgren-Lawrence.  †Separately for all model building approaches.  §Relates to the analysis code, for example, any data cleaning, feature engineering, model building, and evaluation.  ¶Relates to the code to implement the model to get estimates of risk for a new individual. | | | | |

| **Supplementary Table 3. Optimal hyperparameter configuration for the definitive Elastic Net Logistic Regression (EN-LR) pipelines in the CG cohort following feature selection.** | | | |
| --- | --- | --- | --- |
| **Pathway** | **Final Features** | **L1 ratio** | **Regularization (C)** |
| OAI-Complete | 20 | 0.1 | 0.0113 |
| Harmonized | 19 | 0.1 | 0.0113 |
| Hyperparameter values obtained during the final fine-tuning phase (re-optimization) executed exclusively on the cross-validation training folds after Permutation Feature Importance (PFI) dimensionality reduction. L1 Ratio: Defines the Elastic Net mixing parameter, where 0 corresponds to an L2 penalty (Ridge), 1 to an L1 penalty (Lasso), and values in between represent a combination of bot. C (Regularization): Represents the inverse of regularization strength; smaller values specify stronger penalization to prevent overfitting in the remaining feature space. Across models, the algorithm was configured with class_weight='balanced' to manage class imbalance, the saga solver to support Elastic Net penalties, and max_iter=50,000 to allow for maximum coefficient stabilization given the high-dimensional nature of the data. CG denotes Clinical-Genetic; EN-LR, Elastic Net Logistic Regression; and OAI, Osteoarthritis Initiative. | | | |

| **Supplementary Table 4. External calibration performance of the harmonized predictive pipelines of the Clinical-Genetic cohort in the PROCOAC cohort, before and after logistic recalibration.** | | | |
| --- | --- | --- | --- |
| **Pipeline Stage** | **Intercept (Calibration-in-the-large)** | **Calibration Slope** | **O:E Ratio [95% CI]** |
| Original (Uncalibrated) | -0.391 | 0.275 | 1.480 [1.376 – 1.578] |
| Updated (Recalibrated) | 0 | 1.000 | 1.000 [0.921 – 1.083] |
| Calibration metrics of the fully frozen harmonized pipeline evaluated on the external PROCOAC cohort. Original: Uncalibrated predictions reflecting direct geographical translation. Updated: Probabilities adjusted via post-hoc logistic recalibration (Platt scaling). Ideal calibration corresponds to an intercept of 0, a slope of 1, and an O:E ratio of 1. 95% Confidence Intervals (CI) for the O:E ratio were derived via non-parametric cluster bootstrapping (1,000 iterations). CG denotes Clinical-Genetic; CI, Confidence Interval; O:E, Observed-to-Expected; OAI, Osteoarthritis Initiative; and PROCOAC, Prospective Cohort of Osteoarthritis from A Coruña. | | | |

| **Supplementary Table 5. Optimal operational decision thresholds derived from internal cross-validation across all evaluated predictive pipelines.** | | |
| --- | --- | --- |
| **Feature Pathway** | **Sensitive Screening Threshold (F1)** | **High-Specificity Threshold (F0.5)** |
| OAI-Complete | 0.233 | 0.406 |
| Harmonized | 0.259 | 0.408 |
| Thresholds were strictly derived from out-of-fold training predictions in the OAI discovery cohort to prevent optimization bias. The Sensitive Screening Threshold was calculated by maximizing the F1-Score to balance sensitivity and precision. The High-Specificity Threshold was calculated by maximizing the F0.5-Score, weighting precision twice as heavily as sensitivity. CG denotes Clinical-Genetic; CGP and OAI, Osteoarthritis Initiative. | | |

| **Supplementary Table 6. Proposed risk categories for clinical risk stratification of rapid pain progression in knee OA.** | | | |
| --- | --- | --- | --- |
| **Risk category** | **Recalibrated Predicted 24-month risk** | **Clinical meaning** | **Suggested management** |
| Low risk | < 0.26 | Low predicted probability of RPPOA. At this threshold, the model showed a high negative predictive value, indicating that rapid pain progression over 24 months is relatively unlikely, although not completely excluded. | Continue standard knee OA management and routine follow-up. Immediate advanced phenotyping or MRI should not be systematically prioritized based only on model output, but may still be considered if atypical features, marked clinical-radiographic discordance, persistent effusion, or alternative/concomitant diagnoses are suspected. |
| Intermediate risk | 0.26 ‐ < 0.40 | Indeterminate risk zone. RPPOA is not confidently ruled out, but predicted risk does not reach the high-specificity stratum. Clinical evolution and contextual factors remain important for decision-making. | Optimize conservative treatment and reassess within 3–6 months. Consider closer monitoring, repeat clinical evaluation, and advanced phenotyping or MRI if symptoms worsen, pain becomes disproportionate to radiographic findings, inflammatory signs persist, or trial prescreening is clinically relevant. |
| High risk | ≥ 0.40 | High predicted probability of RPPOA. This stratum is enriched for patients likely to experience rapid pain progression and represents a high-specificity profile suitable for downstream clinical or research prioritization. | Prioritize closer monitoring and advanced phenotyping. MRI may be considered when clinically appropriate to characterize pain-relevant structural or inflammatory phenotypes. These patients may also be suitable candidates for clinical trial prescreening or risk-adapted therapeutic strategies. |
| The lower cut-off, 0.26, corresponds to the sensitive screening profile optimized via the F1-score, whereas the upper cut-off, 0.40, corresponds to the high-specificity profile optimized via the F0.5-score. These thresholds are proposed as operational risk-stratification cut-offs derived from the recalibrated model and should not be interpreted as definitive clinical decision rules. MRI assessment, trial referral, or treatment intensification should always be integrated with the full clinical context. Prospective implementation studies are required before routine clinical use. MRI denotes magnetic resonance imaging; OA, osteoarthritis; and RPPOA, rapid pain progression phenotype in osteoarthritis. | | | |
