## Supplemental_methods for "Risk stratification for the rapid pain progression phenotype in knee osteoarthritis using interpretable multimodal machine learning: Development in the Osteoarthritis Initiative and external evaluation in the Prospective Cohort of Osteoarthritis from A Coruña"

**Supplementary Methods**

Supplementary Methods 1. Genomic Data Processing and Variant Selection.

**Genotyping:** In the discovery cohort (OAI), genotyping data were obtained from dbGAP (accession phs000955.v1.p1) yielding a final dataset of 2,946 Caucasian individuals comprising 7,762,204 single nucleotide polymorphisms (SNPs) after processing. For the external evaluation cohort (PROCOAC), independent genotyping was conducted by deCODE genetics using the Illumina GSA chip on an initial sample of 1,511 participants. This cohort underwent systematic reduction, excluding patients lacking the complete 24-month longitudinal clinical follow-up required to accurately compute the rapid pain progression (RPP) phenotype. This resulted in a final PROCOAC analytical dataset of 582 individuals comprising 7,738,208 SNPs.

**Quality Control (QC):** To ensure strict cross-cohort consistency, PROCOAC samples underwent the exact QC pipeline established for the OAI discovery cohort ^1^. Quality control was conducted using PLINK 2.0 ^2^. Initially, variants with low call rates, duplicates, and monomorphic SNPs were removed. Further variant-level filtering excluded SNPs with a minor allele frequency (MAF) < 0.05, Hardy-Weinberg equilibrium (p < 1×10⁻⁶) and > 2% missingness. At the sample level, individuals with > 3% missing genotypes or identified as duplicate samples were excluded. To ensure population homogeneity and data integrity, heterozygosity outliers (> ±4 SD from the mean) and ancestry outliers (> ±8 SD from the European cluster in the 1000 Genomes reference PCA space)^3^ were systematically removed.

**Genetic Imputation and Lead Variant Extraction:** Following QC, VCF files were imputed on the Michigan Imputation Server utilizing Minimac4 ^4^. Only variants demonstrating high imputation quality (R^2^ > 0.8) were retained for downstream analysis. For the machine learning predictive pipelines, our objective was to leverage previously validated genetic signals. We extracted genomic loci demonstrating suggestive association thresholds with RPP (p < $5 \times{10}^{-6}$) from our prior genome-wide analyses. To reduce the genetic feature space, mitigate multicollinearity, and ensure the mathematical independence of predictors, linkage disequilibrium (LD) clumping was applied via PLINK 2.0 ($r^{2}$ > 0.2 in the study population). Within each defined LD cluster, the SNP exhibiting the lowest p-value was designated as the representative index variant. Only these lead variants were incorporated into the feature space of the machine learning algorithms.

Supplementary methods 2. Serum Protein Quantification

To evaluate the potential predictive value of molecular biomarkers, a targeted panel of 11 serum proteins previously implicated in joint metabolism, inflammation, and osteoarthritis pathogenesis was incorporated into the dataset ^5–7^. The panel comprised: Cartilage Oligomeric Matrix Protein (COMP), Chitinase-3-like protein 1 (CHI3L1), Alpha-2-HS-glycoprotein (FETUA), Retinol-binding protein 4 (RET4), Thrombospondin-1 (TSP1), Serum amyloid P-component (SAMP), Apolipoprotein A-I (APOA1), Apolipoprotein A-IV (APOA4), Zinc-alpha-2-glycoprotein (ZA2G), Alpha-2-antiplasmin (A2AP), and Inter-alpha-trypsin inhibitor heavy chain H1 (ITIH1).

Quantification was performed by enzyme-linked immunosorbent assay (ELISA) in the case of ITIH1, while the remaining proteins were measured using antibody suspension bead arrays. COMP, CHI3L1, FETUA, SAMP, TSP1, and APOA1 were quantified using monoplex assays, whereas a multiplex assay was employed for the simultaneous quantification of APOA4, ZA2G, and A2AP. All analyses were carried out strictly according to the manufacturers’ instructions.

Supplementary methods 3. Data Acquisition and Preprocessing

**PROCOAC Cohort Description and Inclusion Criteria:** The Prospective Cohort of Osteoarthritis from A Coruña (PROCOAC) is an independent, longitudinal European observational registry with bi-annual follow-up, established in 2006. Inclusion criteria for the PROCOAC cohort required patients to meet one of the following: i) patients from external consultations with hand pain and diagnosed with hand OA following the American College of Rheumatology (ACR) criteria ^8^; ii) patients with knee pain previously diagnosed with radiographic knee OA following ACR criteria ^9^**;** and iii) patients with hip pain previously diagnosed with radiographic hip OA following ACR criteria ^10^**.**

**Longitudinal Data Extraction:** In the OAI discovery cohort, the 10-year follow-up period was deliberately truncated to preserve data density. The 24-month overlapping longitudinal instances were constructed utilizing data from the following specific timepoints: baseline (V00), 12-month (V01), 24-month (V03), 36-month (V05), 48-month (V06), 72-month (V08), 96-month (V10), and 120-month (V12) visits. Because time between visits in the PROCOAC cohort exhibited natural clinical variance, a dynamic time-window extraction was implemented. Transitions between consecutive visits were retained as valid overlapping instances if the elapsed time between assessments fell within a 20- to 30-month interval, ensuring temporal comparability with the OAI cohort.

**Environment and Data Integration:** Data preparation and preprocessing were conducted utilizing R version 4.5.1 ("Great Square Root") within the RStudio environment ^11^. The initial dataset integrated the official OAI "All Clinical" and "Enrollees" longitudinal datasets covering the 120-month follow-up period, initially comprising 8,308 clinical and 60 enrollee variables. Administrative and technical metadata were systematically excluded. Radiographic severity (Kellgren-Lawrence grading) was previously assessed by radiologists and/or trained rheumatologists from the original OAI and PROCOAC cohorts, who were blinded to the longitudinal clinical progression of the patients. This clinical matrix was subsequently enriched with multi-omic data, specifically: quantitative proteomic biomarkers, selected participant genotypes from prior genome-wide association studies (GWAS), and targeted mitochondrial variants.

**Bilateral Consolidation and QC:** To address the side-specific nature of osteoarthritis data, a bilateral feature consolidation step was executed. Left- and right-side knee variables were paired and aggregated to retain the value indicating greater severity (maximum or minimum, depending on the specific variable's clinical directionality). This approach effectively captured the "worst-knee" presentation per longitudinal instance, maximizing information retention. Prior to imputation, features were systematically classified into continuous, ordinal, or categorical variables based on nomenclature and statistical distribution. Strict QC filters were applied: features exhibiting >50% missing values or near-zero variance were removed from the feature space, and longitudinal instances with >40% missing data were excluded.

**Data Splitting:** To strictly prevent data leakage and optimism bias, the dataset was partitioned into training (80%) and testing (20%) sets based exclusively on unique patient IDs. This strategy guaranteed that all overlapping longitudinal instances belonging to a single individual were assigned to the same partition, preserving the natural balance of rapid pain progression cases across both sets.

**Longitudinal Imputation:** Missing data were addressed via a two-stage longitudinal strategy: (1) Intra-subject, where missing values within individual patient trajectories were resolved using linear interpolation for continuous numeric features and forward-filling (last-known-value) for ordinal variables and (2) Global, where remaining unresolvable missing values were imputed using the global median (for continuous/ordinal variables) or mode (for categorical variables). All imputation parameters (medians and modes) were derived strictly from the isolated 80% training set and subsequently applied to the testing set.

Supplementary methods 4. Cohort Stratification and Cross-Cohort Feature Harmonization

**Analytical Cohort Stratification:** To maximize statistical power while accounting for the differential availability of specific multi-omic data layers, the study population was formally stratified into two overlapping analytical datasets. First, the Clinical-Genetic (CG) cohort comprised all eligible Caucasian participants with clinical records and matched genomic data. Second, the Clinical-Genetic-Proteomic (CGP) cohort was defined as a nested subset comprising individuals for whom quantitative proteomic profiling was additionally available. This stratification allowed for the evaluation of the prognostic value of different molecular biomarkers.

**Dual-Pathway Experimental Design and Harmonization Filter:** The first pathway ("OAI-Complete") utilized the maximal available feature space in the discovery cohort after preprocessing, yielding an initial space of 354 features for the CG cohort and 367 for the CGP cohort. For the second pathway ("Harmonized") the final feature space was restricted exclusively to variables routinely collected in the European PROCOAC cohort. This harmonization was achieved through an iterative backward elimination process. Features identified as highly predictive by the model in the OAI discovery cohort were cross-referenced against the PROCOAC data dictionary. If a selected OAI feature, e.g., specific Knee Injury and Osteoarthritis Outcome Score (KOOS) subscales, lacked a direct clinical equivalent in PROCOAC, it was systematically omitted from the analytical space, and the modelling pipeline was retrained from scratch. This iterative pruning cycle continued until the algorithm's selected feature signature comprised exclusively variables available in both cohorts, yielding a final harmonized initial space of 159 variables for the CG cohort and 164 for the CGP cohort (prior to redundancy filtering).

Supplementary methods 5. Algorithm Benchmarking, Feature Selection, and Pipeline Assembly

**Computational Environment and Preprocessing:** Machine learning analyses were conducted in Python 3.12.7 ^12^. Prior to modelling, multicollinearity was addressed by excluding highly correlated features based on data-specific thresholds: >0.80 for continuous (Pearson) and ordinal (Spearman) variables, and >0.85 for categorical features (Cramer’s V). This redundancy filtering was applied prior to any data transformation in the OAI-Complete dataset, and immediately post-harmonization in the Harmonized dataset. Model development was fully encapsulated within scikit-learn pipelines to guarantee methodological rigor and strictly prevent data leakage during cross-validation. The preprocessing stage consisted of a numerical transformer applying Z-score standardization and a categorical transformer employing One-Hot Encoding. The encoder was forced to learn a fixed feature space strictly from the OAI training set.

**Algorithm Benchmarking and Class Imbalance Strategy:** To manage class imbalance of the rapid pain progression phenotype during model training, cost-sensitive learning (class weighting) was applied to the tree-based models, SVM-RBF, and EN-LR. Conversely, the Synthetic Minority Over-sampling Technique (SMOTE)^13^ was applied to the MLP exclusively within the training folds. A Randomized Search, with a 5x5 Repeated Stratified Group K-Fold cross-validation scheme, was employed to explore the hyperparameter space of each algorithm. Patient IDs were utilized as the grouping variable across all folds to strictly prevent intra-subject data leakage. Optimization prioritized PR-AUC, as it provides a more robust performance estimation than the Area Under the Receiver Operating Characteristic Curve (ROC-AUC) in highly imbalanced datasets ^14^. Complete search grids and distribution ranges are detailed in **Supplementary Table 1.**

**Feature Pruning and Pipeline Re-optimization:** Following the selection of EN-LR ^15^ as the optimal algorithm, dimensionality reduction was performed across both pathways using Permutation Feature Importance (PFI) ^16^. To avoid computational bottlenecks, PFI was calculated across the 80% training set to derive a fixed, reduced feature space prior to hyperparameter cross-validation. While establishing a global feature signature outside the internal CV folds may introduce optimism bias within the internal tuning scores, protection against data leakage was guaranteed during model evaluation. The 20% hold-out test set and the external PROCOAC cohort were excluded from all feature selection procedures, ensuring that final performance metrics remained completely unbiased. Features were ranked based on their PFI score (PR-AUC drop), isolating critical predictors while discarding noise (importance score threshold > 0.001). Following feature reduction, a final hyperparameter re-optimization (specifically, penalization strength and L1 ratio) was performed on the reduced datasets. The preprocessor was forced to re-learn scaling and encoding parameters from the cross-validation training folds.

**Nested Cross-Validation for Calibration and Threshold Optimization:** Isotonic Regression ^17,18^ via nested cross-validation was applied to calibrate the final probabilities and threshold selection. First, an inner calibration loop utilized CalibratedClassifierCV with non-parametric Isotonic Regression (method=’Isotonic’) and an internal 5-fold scheme (cv=5). The definitive models were then deployed as end-to-end pipelines comprising the preprocessor, a custom feature filter, and the Calibrated Classifier ensemble. Second, an outer validation loop (5-fold StratifiedGroupKFold scheme) was implemented. This loop iteratively cloned the complete calibrated pipeline, with different seeds, trained it on 80% of the patient clusters, and generated out-of-fold (OOF) probabilities for the remaining 20% unseen patients. Patient-level grouping prevented intra-subject data leakage. Using the accumulated OOF precision-recall curve, two operational thresholds were defined: (1) The sensitive screening threshold (maximizing the F1-score) to balance sensitivity and precision and (2) the high-specificity enrichment threshold (maximizing the F0.5-score, weighting precision twice as heavily as recall) to minimize false positives.

Supplementary methods 6. Statistical Evaluation, Hypothesis Testing, and Model Interpretability

**Non-Parametric Cluster Bootstrapping:** To evaluate prediction uncertainty and calculate robust 95% confidence intervals for all discriminative and calibration metrics, a non-parametric cluster bootstrapping approach (1,000 iterations) was implemented^19,20^. Rather than resampling individual longitudinal instances (which would violate the assumption of independence and artificially narrow the confidence intervals) resampling was conducted at the patient level (cluster). During each iteration, patient IDs were sampled with replacement, and the sample was reconstructed by concatenating all available longitudinal instances belonging exclusively to the selected patients^20^. This cluster-bootstrapping approach was also applied to the Decision Curve Analysis (DCA) to calculate 95% confidence intervals for the Net Benefit ^21^.

**Generalizability Testing:** To quantify the "cost of generalizability" between the OAI-Complete and Harmonized pathways, statistical comparisons of discriminative performance (ROC-AUC and PR-AUC) were conducted using a resampling non-parametric cluster-paired permutation test (5,000 iterations) ^20,22^. This technique was selected to account for the correlated nature of predictions derived from the overlapping longitudinal instances of the same individuals within the hold-out test set.

**Model Interpretability (SHAP**): Global and local model interpretability were derived utilizing SHapley Additive exPlanations (SHAP) ^23^. Given the selection of the penalized linear model (EN-LR) as the definitive pipeline, the algorithm's underlying mechanistic logic was quantified via shap.LinearExplainer.

Supplementary methods 7. External Evaluation and Model Updating

**External Calibration and Logistic Recalibration:** Model performance in the external cohort was evaluated before (raw) and after updating. Calibration was quantified using the Brier Score, calibration curves, calibration slope, and the Observed-to-Expected (O:E) ratio. Recognizing the baseline prevalence shift in rapid pain progressors between the discovery (OAI: ~10%) and evaluation (PROCOAC: ~28%) cohorts, a Model Updating procedure was conducted following TRIPOD+AI guidelines ^24^. To correct the systematic underestimation driven by this epidemiological shift, a post-hoc logistic recalibration (intercept and slope updating) was applied ^19,25,26^. This procedure mathematically adjusted the model’s baseline risk output to the PROCOAC clinical context without re-estimating the individual feature coefficients learned during internal training, preserving their relative prognostic value and preventing external overfitting.
