## Supplemental_figures for "Risk stratification for the rapid pain progression phenotype in knee osteoarthritis using interpretable multimodal machine learning: Development in the Osteoarthritis Initiative and external evaluation in the Prospective Cohort of Osteoarthritis from A Coruña"

**Supplementary figures**

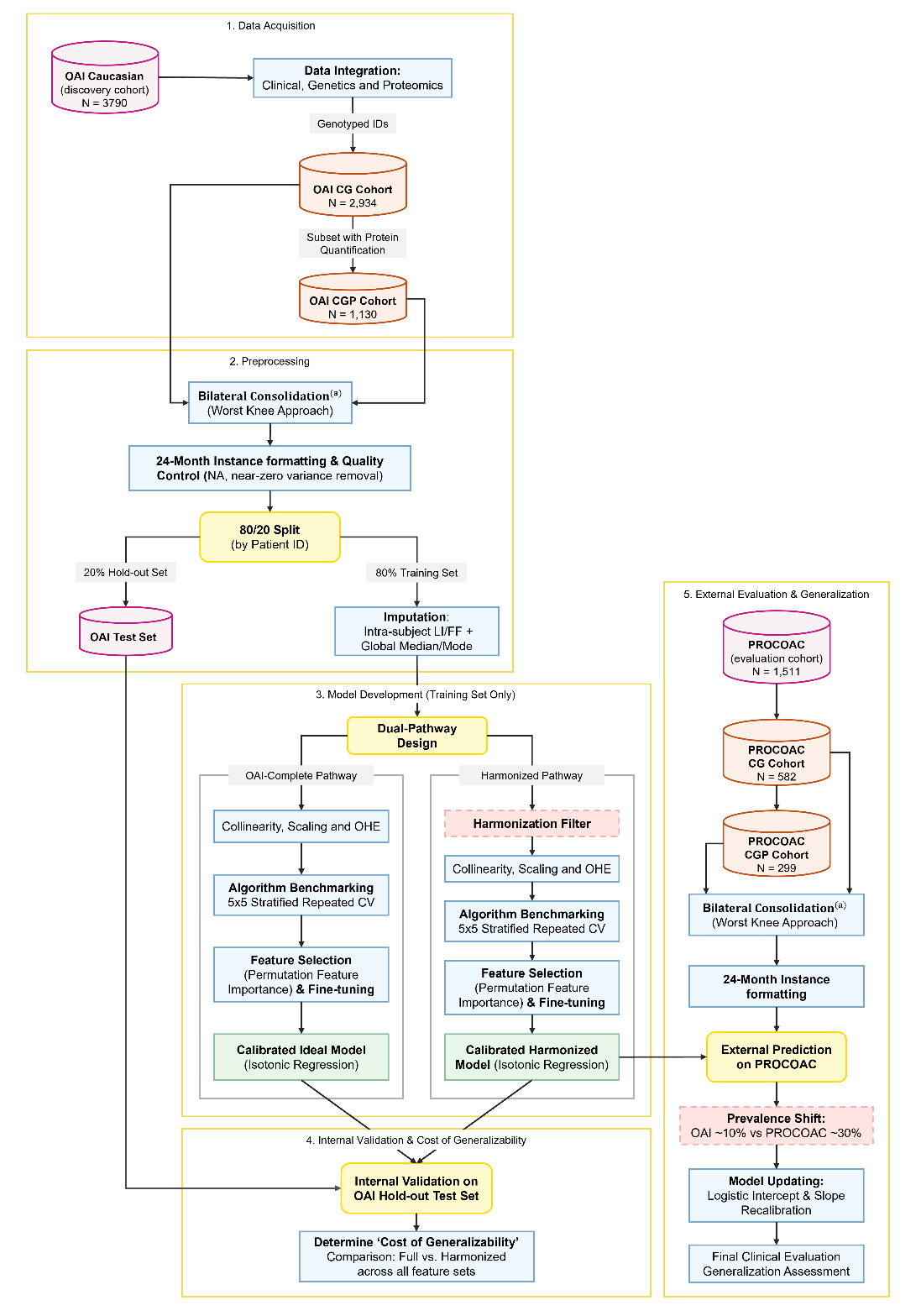

**Supplementary Figure 1. Methodological workflow of the multi-cohort study.** The analytical pipeline encompasses five distinct phases: (1) Data Acquisition and integration of the OAI discovery cohort with molecular datasets; (2) Preprocessing, including bilateral ('worst-knee') consolidation, quality control, and longitudinal imputation; (3) Model Development through a dual-pathway experimental design (OAI-Complete vs. Harmonized), applying a specific harmonization filter to the latter, followed by algorithm benchmarking, feature selection, fine-tuning, and calibration; (4) Internal Validation on the independent OAI hold-out test set; and (5) External evaluation and transportability to the European PROCOAC cohort, applying logistic recalibration to address prevalence shifts. ^(a)^ All subsequent steps were executed independently for the CG Cohort and the CGP cohort. CG denotes Clinical-Genetic; CGP, Clinical-Genetic-Proteomic; CV, Cross-Validation; FF, Forward-filling; LI, Linear Interpolation; NA, Not Available (missing value); OAI, Osteoarthritis Initiative; OHE, One-Hot Encoding; and PROCOAC, Prospective Cohort of Osteoarthritis from A Coruña.

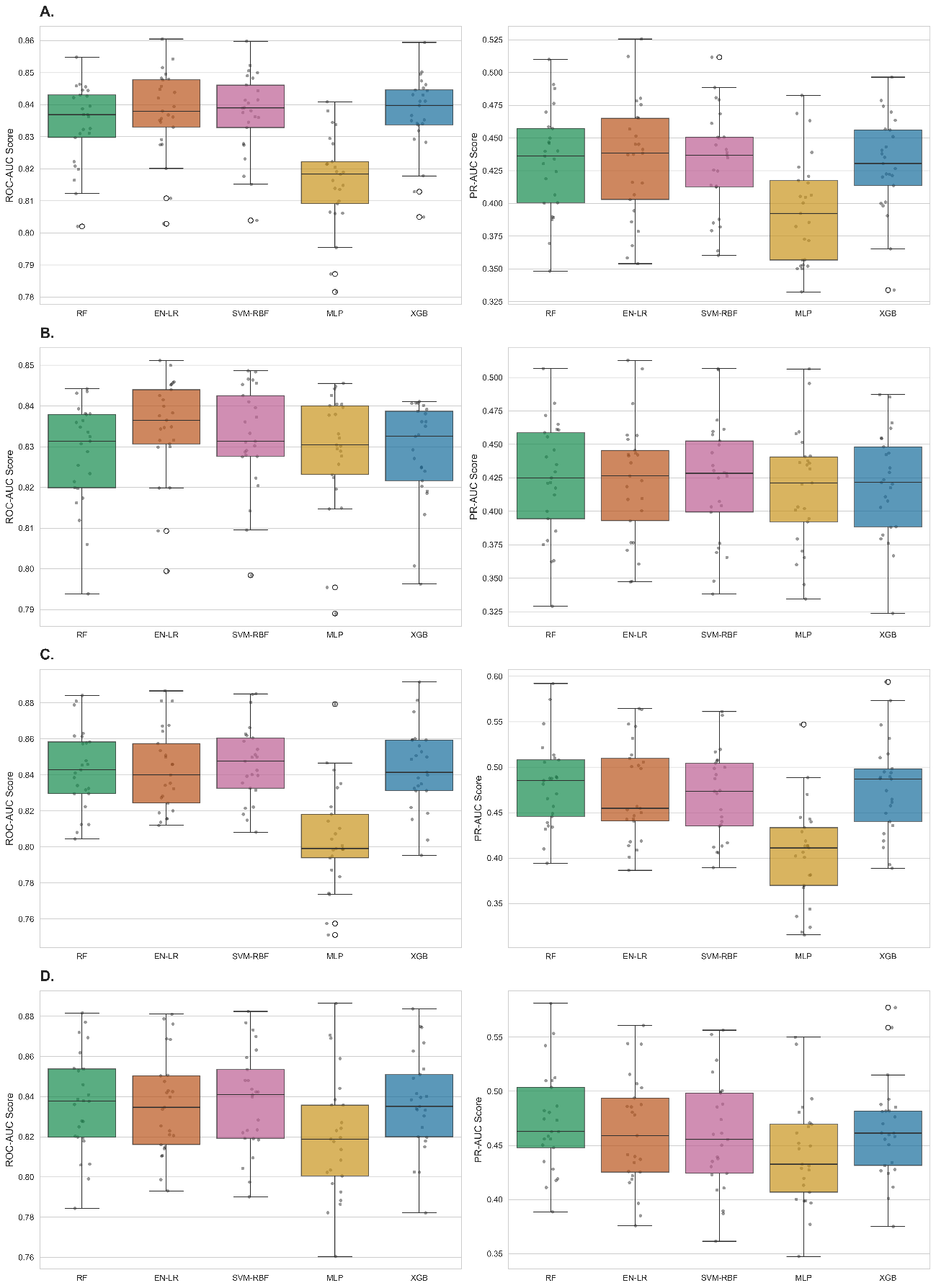

**Supplementary Figure 2. Cross-validation stability and predictive robustness across the evaluated machine learning algorithms.** Boxplots illustrate the distribution of the 25 evaluation scores (ROC-AUC on the left and PR-AUC on the right) obtained during the 5x5 Repeated Stratified Group K-Fold Cross-Validation phase on the training sets. Each panel represents a modelling scenario: (A) Clinical-Genetic (CG) cohort employing the OAI-Complete feature space; (B) CG cohort employing the harmonized feature space; (C) Clinical-Genetic-Proteomic (CGP) cohort employing the OAI-Complete feature space; and (D) CGP cohort employing the harmonized feature space. Black dots overlaid on the boxplots represent the individual metric scores for each validation fold. Across all high-dimensional setups, the Elastic Net Logistic Regression (EN-LR) consistently demonstrated high discriminative power alongside minimal inter-fold variance compared to the non-linear alternatives. CG denotes Clinical-Genetic; CGP, Clinical-Genetic-Proteomic; EN-LR, Elastic Net Logistic Regression; MLP, Multilayer Perceptron; PR-AUC, Precision-Recall Area Under the Curve; RF, Random Forest; ROC-AUC, Receiver Operating Characteristic Area Under the Curve; SVM-RBF, Support Vector Machine with Radial Basis Function kernel; and XGB, eXtreme Gradient Boosting.

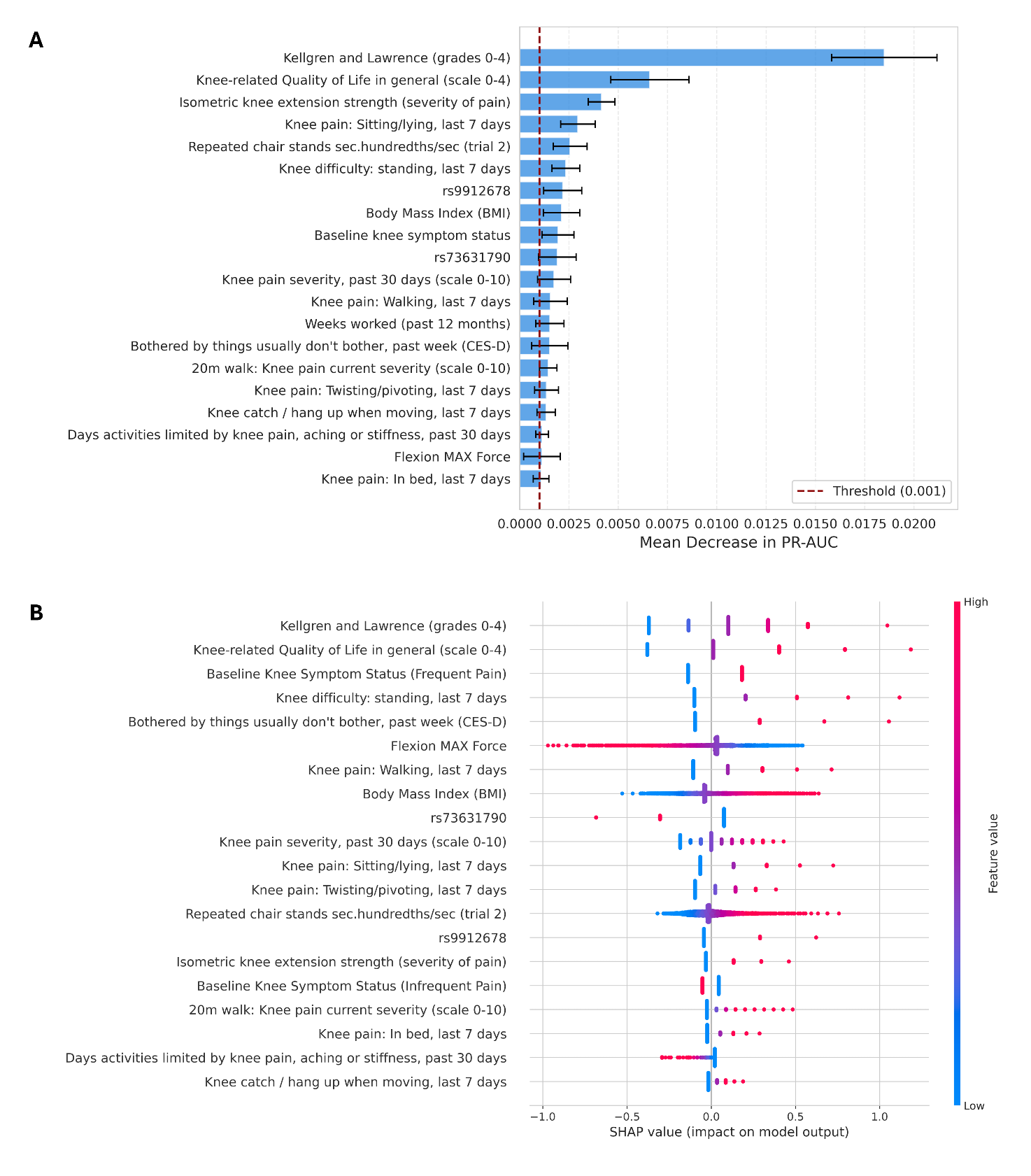

**Supplementary Figure 3. Global interpretability of the OAI-Complete model in the Clinical-Genetic (CG) Cohort.** (A) Permutation feature importance ranking. Variables are sorted by their mean impact on the model's predictive performance (PR-AUC decrement) when permuted. Error bars represent the 95% confidence intervals across repeated permutations. (B) SHapley Additive exPlanations (SHAP) summary dot plot evaluating the directionality and magnitude of each feature's contribution to the prediction of rapid pain progression. Each dot represents a single patient's hold-out test evaluation. Color indicates the actual feature value (red = high, blue = low), and the position on the x-axis indicates the SHAP value (impact on the probability of rapid pain progression). CES-D denotes Center for Epidemiologic Studies Depression Scale; m, meters; Max, Maximum; OA, Osteoarthritis; SF-12, 12-Item Short Form Health Survey; and WOMAC, Western Ontario and McMaster Universities Osteoarthritis Index.
